# Non surgical procedure related postoperative Complications independently predicts perioperative mortality, in gastrointestinal and Hpb surgeries.- A retrospective Analysis of prospectively maintained data

**DOI:** 10.1101/2020.04.17.20068940

**Authors:** Bhavin Vasavada, Hardik patel

## Abstract

**Aim:** The Aim of the study was to evaluate relationship between non surgical procedure related complication and 30 days mortality.

**Material and Methods:** All gastrointestinal and hepatobiliary procedures performed in last 3 years have been evaluated retrospectively. Non surgical procedure related postoperative complications were defined as perioperative complications non related to surgical procedures or techniques and related to patients’ physiological health or comorbidities (e.g acute kidney injury, ARDS, acute respiratory failure, pre existing sepsis, etc.), Surgical related complications were defined as perioperative complications related to surgical procedures or techniques (e.g. bleeding, leaks, sepsis due to leaks etc.). Factors affecting 30 days mortality and morbidity were analysed using univariate and multivariate analysis. Chi square test was used for categorical values, Mann Whitney U test was used for numerical values. Multivariate logistic regression analysis was used for multivariate analysis. Statistical analysis was used suing SPSS version 21.

**Results:** Total 325 major hepatobiliary and pancreatic surgery was done in our institute in last 2 years. 30 days overall mortality rate was 6.4%. In univariate analysis mortality was significantly associated with nonsurgical procedure related complications. (p < 0.0001). Surgical complications were not associated with mortality. On univariate analysis other factors associated with mortality were emergency surgeries, high CDC grade of surgery, higher ASA grades, increase operative duration, increased blood product requirements. However on multivariate analysis only nonsurgical procedure related postoperative complications independently predicted mortality. (p=0.001).

**Conclusions:** Non surgical procedure related post operative complications (Physiological) is strongly associated with 30 days mortality, suggesting improved perioperative care can help to reduce post operative mortality.

## Introduction

Surgical Complications are major cause of mortality and morbidity. [1] Surgical complications can be as high as 30% in some group of patients. [2,3]. Surgical complications generally consist of two type of complications as per our knowledge. One is technique or surgical procedure related e.g. bleeding, anastomotic leaks. The other is non surgical technique related complications or complications due to change in physiology due to surgical stress, for example ARDS, Acute kidney Injury, Post operative Acute left ventricular failure or Post operative acute delirium. In non surgical technical related complication prehabilitation can work. [4]. We hypothesised that non surgical technique related complications or what we call physiological complications are much more common than surgical technique related or surgical complications.

## Aim

The Aim of the study was to evaluate relationship between non surgical procedure related complication and 30 days mortality.

## Material and Methods

All gastrointestinal and hepatobiliary procedures performed in last 3 years have been evaluated retrospectively.

Nonsurgical procedure related postoperative complications were defined as perioperative complications non related to surgical procedures or techniques and related to patients’ physiological health or comorbidities (e.g acute kidney injury, ARDS, acute respiratory failure, pre existing sepsis, etc.).

Surgical related complications were defined as perioperative complications related to surgical procedures or techniques (e.g., bleeding, leaks, sepsis due to leaks, etc.). Factors affecting 30 days mortality were analysed using univariate and multivariate analysis.

### Statistics

Fisher t test was used for categorical values, Mann Whitney U test was used for numerical values. Multivariate logistic regression analysis was used for multivariate analysis. Statistical analysis was used suing SPSS version 21.

## RESULTS

Total 325 major hepatobiliary and pancreatic surgery was done in our institute in last 2 years. 30 days overall mortality rate was 6.4%.

In univariate analysis mortality was significantly associated with nonsurgical procedure related complications. (p < 0.0001). Surgical complications were not associated with mortality. (p=0.235)

On univariate analysis other factors associated with mortality were emergency surgeries, high CDC grade of surgery, higher ASA grades, increase operative duration, increased blood product requirements. [Table 1]

**Table: 1.**
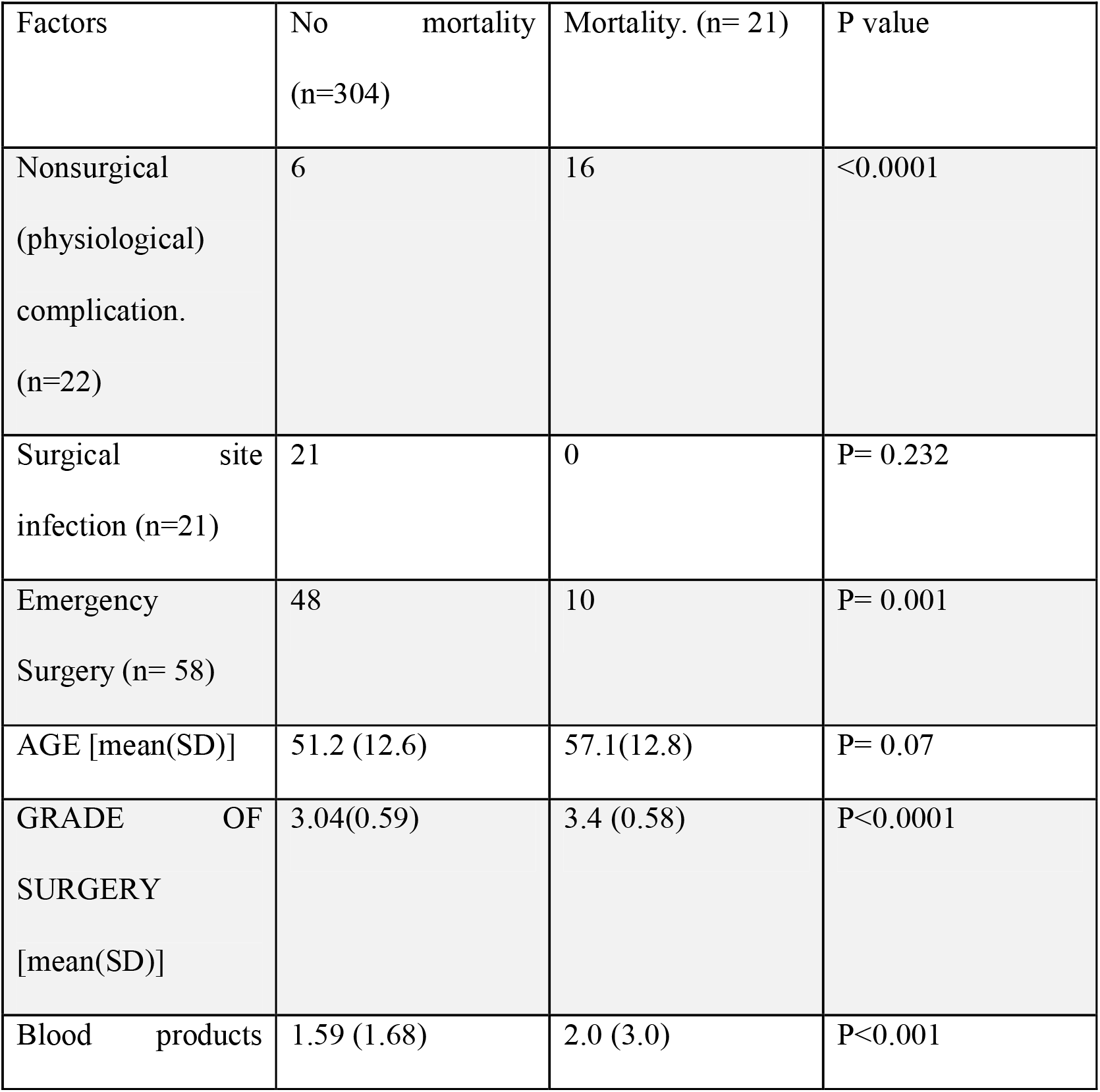
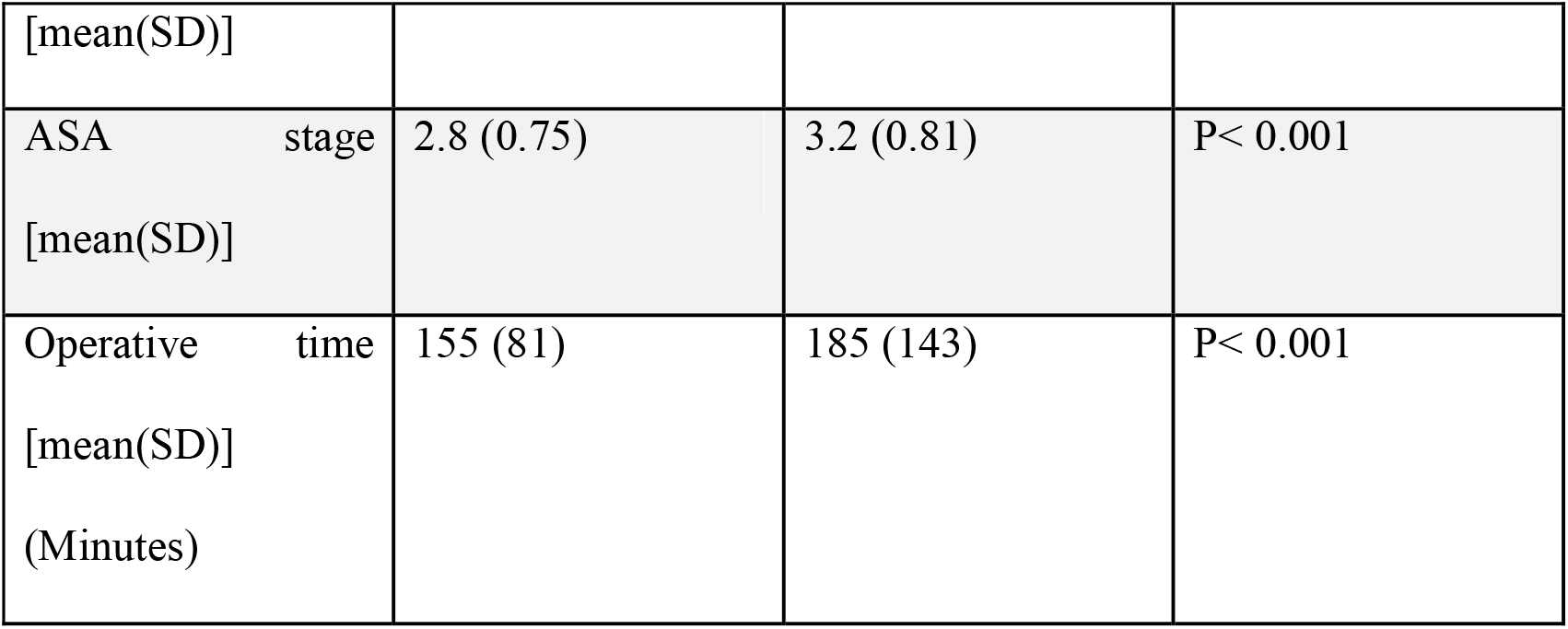
Univariate analysis for 30 days mortality

However on multivariate analysis only nonsurgical procedure related postoperative complications independently predicted mortality. (p <0.001). [Table 2] Odds ratio was 0.007 with 95% confidence interval 0.001 to 0.038.

**Table 2.**
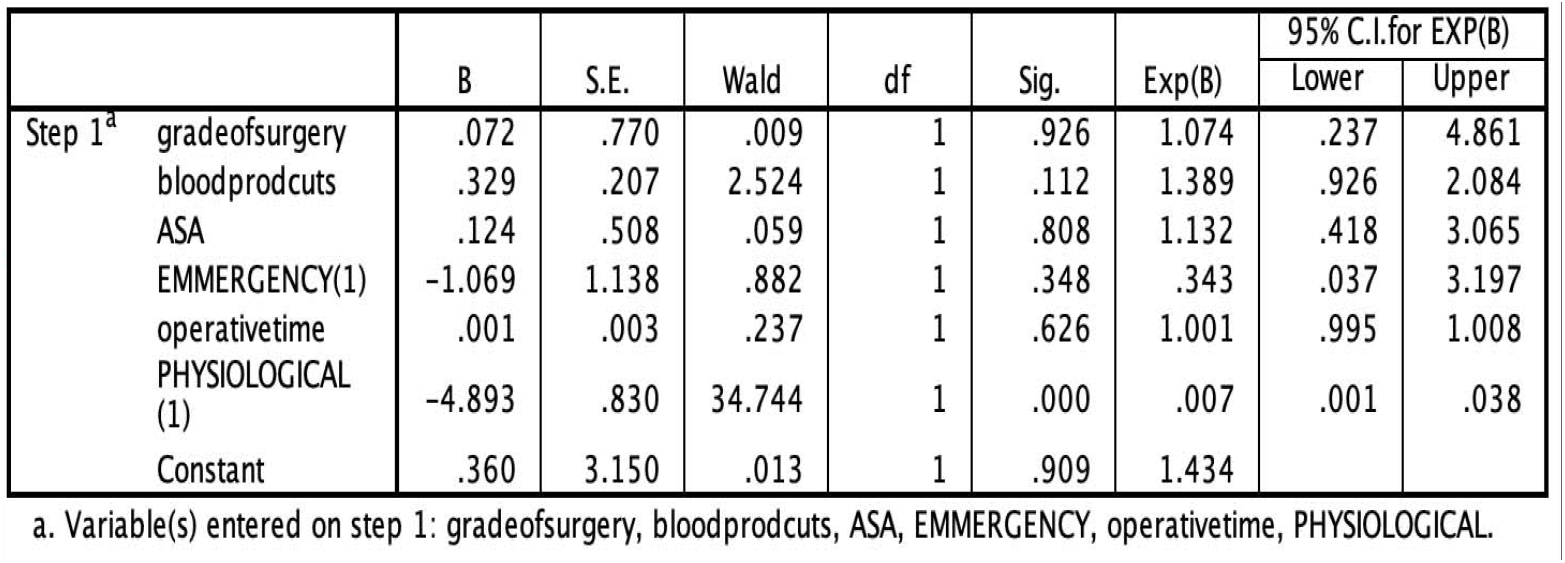
multivariate logistic regression

Table :1 Univariate analysis for 30 days mortality
| Table 2. multivariate logistic regression |  |  |  |  |  |  |  |  |
| --- | --- | --- | --- | --- | --- | --- | --- | --- |
|  | B | S.E. | Wald | df | Sig. | Exp(B) | 95% C.I. for EXP(B) |  |
|  |  |  |  |  |  |  | Lower | Upper |
| Step 1<sup>a</sup> |  |  |  |  |  |  |  |  |
| gradeofsurgery | .072 | .770 | .009 | 1 | .926 | 1.074 | .237 | 4.861 |
| bloodprodcuts | .329 | .207 | 2.524 | 1 | .112 | 1.389 | .926 | 2.084 |
| ASA | .124 | .508 | .059 | 1 | .808 | 1.132 | .418 | 3.065 |
| EMMERGENCY(1) | -1.069 | 1.138 | .882 | 1 | .348 | .343 | .037 | 3.197 |
| operativetime | .001 | .003 | .237 | 1 | .626 | 1.001 | .995 | 1.008 |
| PHYSIOLOGICAL (1) | -4.893 | .830 | 34.744 | 1 | .000 | .007 | .001 | .038 |
| Constant | .360 | 3.150 | .013 | 1 | .909 | 1.434 |  |  |

a. Variable(s) entered on step 1: gradeofsurgery, bloodprodcuts, ASA, EMMERGENCY, operativetime, PHYSIOLOGICAL.

Kaplan Meier 30 days survival curve also showed mortality was significantly higher in patients with nonsurgical technique related complications. P value was less than 0.0001 with log rank test. [Figure 1]. However there was no difference in patients’ with surgical technique related complications.

**Figure 1.**
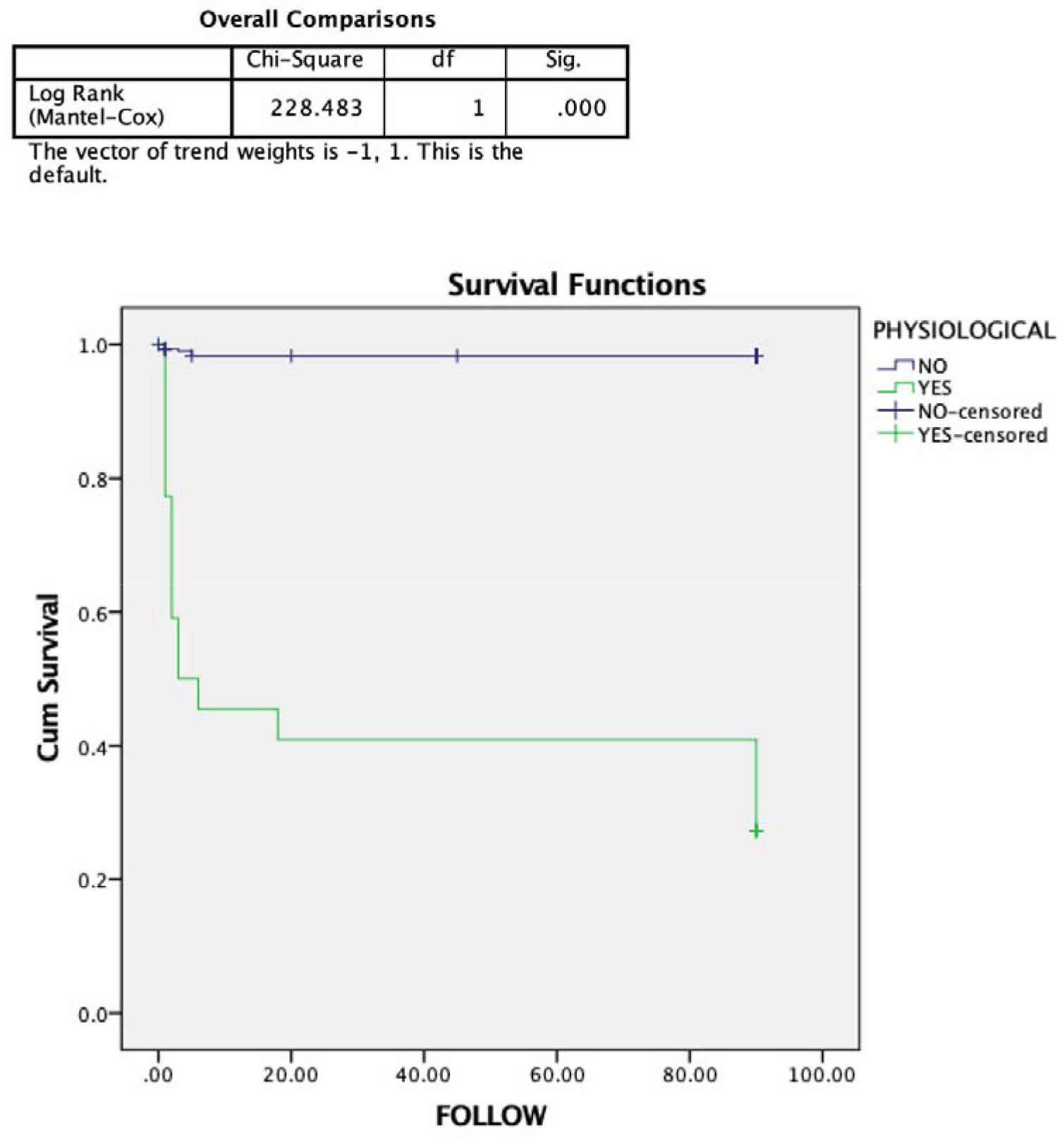
30 day survival comparison with non surgical complication and surgical complication with log rank analysis. (p< 0.0001)

## Discussion

Perioperative mortality is the one the most important problem surgical community is facing. Peri operative mortality range from 0.1% to as high as 27%-30% according to various types of surgeries. [5]. Gastro intestinal and hepatobiliary surgeries are technically demanding procedures and has one of the highest perioperative mortality rates. [6,7,8]

Surgeons are always worried about surgical techniques and surgical techniques related mortalities, how ever a very few studies are done to look at the impact of such complications on perioperative mortality. There are various perioperative complications which are not actually related to surgical techniques, but depend on many factors like patients’ pre operative conditions as well as perioperative anaesthetic course and complications due to that. These complications can include but not limited to acute kidney injury, adult respiratory distress syndrome, post operative delirium, non surgery related sepsis and septic shock. These complications can contribute to mortality significantly. [9.10]

Aim of our study to look for effect of nonsurgical technique related complications and surgical technique related complications on mortality. For gastrointestinal and hepatobiliary surgeries we defined anastomotic leaks, sepsis due to leaks,intraoperative bleeding, iatrogenic injuries to surrounding structure as surgical technique related complications and other complications like acute kidney injury, ards (acute respiratory distress syndrome), sepsis not related to surgical technique etc we defined as non surgical technique related complications of physiological complications.

Surgery related complications were not associated with mortality. On univariate analysis, physiological complications, ASA score, blood product requirements, operative time, CDC grade of surgery [11], and emergency surgery were associated with 30 days mortality. [table 1] Suggesting patient related factors were associated with 30 days mortality rather than surgical technique related complications. Which is also supported by current literature available. [12,13,14,15]

However on multivariate analysis only occurrence of nonsurgical technique related or what we call as physiological complications predicted, survival outcomes. [table 2]. Kaplan Meier survival curves also predicted lower 30 days survival for patients who had physiological complications. [figure 1] but there was no difference in 30 days survival in patient who has surgical technique related complications. [Figure 2]

**Figure 2:**
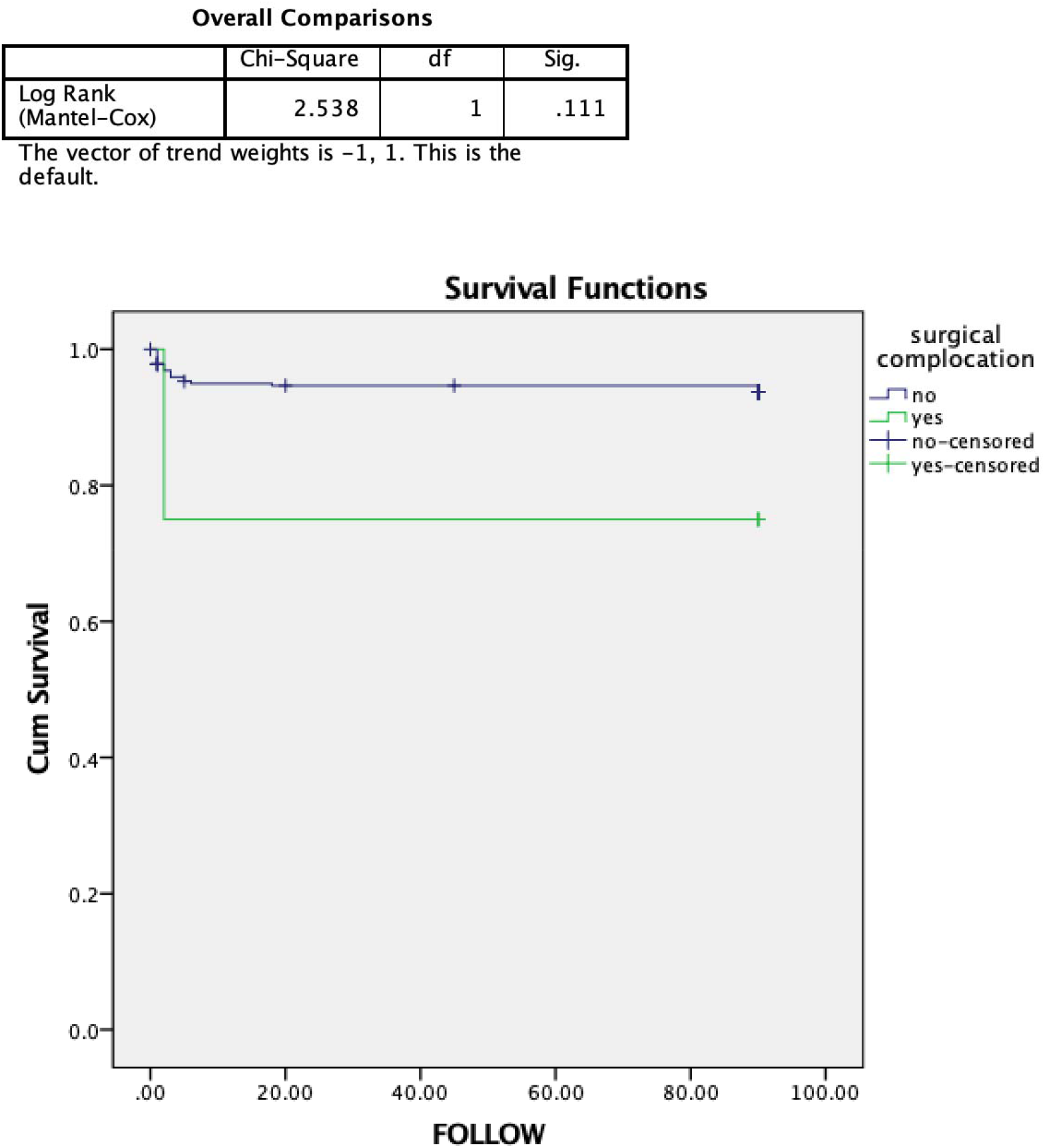
There was no difference in 30 days survival in patient with surgical technique related complications.

We do not want to say that surgical technique related complications are not harmful but our purpose is to point out importance of nonsurgical technique related complications and their impact on surgical mortality. This study also shows importance of critical care management in reducing postoperative mortality. [16,17,18,19].

Being a retrospective study this study has inherent limitations associated with it. Another limitation is its includes wide variety of sub specialities. However we believe these findings will remain same across the specialities.

In conclusion Non surgical procedure related post operative complications (Physiological) is strongly associated with 30 days mortality, suggesting improved perioperative critical care can help to reduce post operative mortality.

## Data Availability

Raw data can be made available on demand.

## Notes

### Competing Interest Statement

The authors have declared no competing interest.

### Funding Statement

no funding obtained.

